# Standardising the structure of routinely collected data for childhood ocular inflammation: a SNOMED-CT mapping project

**DOI:** 10.1101/2023.10.25.23297537

**Authors:** Salomey Kellett, Ameenat Lola Solebo, the Paediatric Ocular Inflammation UNICORN Study Group

## Abstract

**Background and aims:** Multicentre, longitudinal research methods are usually necessary for rare disease research. SNOMED CT (Systematized Nomenclature of Medicine Clinical Terms), the comprehensive and standardized terminology system can be used to enhance the interoperability of data collected across different settings. Childhood uveitis is a rare, blinding disorder, with uncertainties around disease distribution and outcome. To enhance the interoperability of uveitis data, we created a SNOMED CT coded dataset derived from a core clinical dataset.

**Methods:** Data elements were selected from a published list developed through a consensus exercise undertaken by a national disease research group, the United Kingdom’s Paediatric Ocular Inflammatory Group (POIG). Items were organised using a three level priority score, based on the National Institute for Health (NIH) model for common data elements, and grouped using the Heath Level 7 (HL7) standard "Fast Healthcare Interoperability Resources" (FHIR) generic data structure, and then mapped across to the SNOMED CT codes.

**Results:** From the POIG consensus exercise, 160 elements were selected: 89 as high priority items, with 35 as medium and 29 as low priority items. These elements, and response items where appropriate, were grouped into Patient (n= 13 items), Observation (n= 63 items), Condition (n= 20 items), Procedure (n= 44 items), Medication (n= 18 items). There were four items for which a SNOMED CT ID could not be found.

**Conclusion:** Through this mapping activity, using international coding and terminologies, we have created a dataset for childhood onset uveitis care and research. This dataset provides a standardised vocabulary for describing clinical concepts, with a semantic interoperability which will support the exchange of data across different systems, organizations, and international or supranational groups. Future expansion of the dataset will be needed to ensure coverage of international concepts and care structures.

**Author summary:** Through a mapping activity, in which data items from a consensus developed core clinical dataset were mapped across to SNOMED CT terminologies, we have created a dataset for childhood onset uveitis care and research. This dataset provides a standardised vocabulary for describing clinical concepts, with a semantic interoperability which will support the exchange of data across different systems, organizations, and international or supranational groups.

## Introduction

Childhood uveitis comprises a group of individually rare inflammatory disorders which exhibit different clinical ocular and systemic phenotypes,(1) and confer a life-long risk of risk of visual morbidity.(2,3) In up to 25% of children with uveitis, a degree of permanent vision loss will affect at least one eye,(4) and the disease, and attendant risk of further visual loss, often persists into mid-adulthood.(5) Prompt diagnosis and prompt disease control are key in preventing uveitic blindness. Achieving this control necessitates a combination of topical and systemic corticosteroids, systemic disease-modifying immunomodulatory agents, and biologic therapies, with agents typically chosen in a trial and error approach. Alongside the need to personalise therapeutic approach, other evidence gaps in the field include aetiology, with the majority of cases being idiopathic,(6) disease burden, with studies reporting a 20-fold difference in incidence rates,(6–8) and the uncertainty around the determinants of long term positive and negative eye and global health outcomes.(9,10)

Routinely collected clinical data have been harnessed across different disease areas to enable observational research on disease incidence,(11) prognostication,(12) and to support interventional studies.(13) For rare diseases, where multi-centre work is needed to reach the sample sizes sufficient to undertake such research, the re-use of routinely collected clinical data is dependent on the interoperability of data extracted across different sites.(14) Several groups have developed approaches and resources with the aim of defining standardized datasets for the systematic collection of uveitis-related information.(15–17) However, while these represent important advances, more is needed to support interoperability, specifically, the common data elements within the developed datasets must be supported by coding derived from a clearly structured terminology standard.

The SNOMED CT (Systematized Nomenclature of Medicine Clinical Terms) is a comprehensive and standardized terminology system widely used in healthcare and research.(18) To enhance the interoperability of uveitis data, we created a SNOMED CT coded dataset derived from a core clinical dataset, developed in collaboration with the United Kingdom’s Paediatric Ocular Inflammation Group (POIG).(15,19) We present an overview of the dataset and its development process.

## Methods

We undertook the development of a dataset which used an international terminology (SNOMED CT) to support interoperability.

### Selection of data elements

The POIG core clinical dataset for non-infectious uveitis was used as a source of data elements for this mapping exercise. POIG is a multi-disciplinary group formed of clinicians who manage children with complex inflammatory disorders, with a central aim of delivering the evidence base needed to support the care of these children, and supporting the translation of that evidence base into changes in policy and practice. The group developed, through Delphi consensus, as reported elsewhere,(15) a consensus dataset for the routine collection of data for children diagnosed with uveitis. This dataset includes common data items (CDEs) on demographic details, pathways to diagnosis, ocular disease phenotype, activity level and severity scores, the presence, type and severity of co-existent systemic disease, clinical interventions, and laboratory and imaging findings. Those data items which formed the POIG core clinical dataset (having been agreed upon through the consensus exercise) were selected. Other items, which had been suggested within the ‘long-list’ developed by the Delphi group, were also selected (table 1). These were judged to be ‘low priority’ items, versus the ‘high priority’ items comprising the core clinical dataset. Those items which were agreed upon by the majority of the Delphi group, but which had not reached the threshold for formal inclusion in the core clinical dataset (table 1), were coded as ‘moderate priority’. This three level prioritisation scoring was based on the National Institute for Health (NIH) model for common data elements (CDEs) (table 1).(20)

**Table 1.** Prioritisation of data elements.

| Priority | NIH classification(20) | Definition |
| --- | --- | --- |
| High | General core/disease core* | Items included in the core childhood uveitis clinical dataset following consensus decision by Delphi group, ie inclusion supported by >95% |
| Moderate | Supplemental—highly recommended | Items supported by majority of Delphi group (>50%) but not reaching consensus threshold for inclusion in core dataset |
| Low | Supplemental or Exploratory | Items supported by less than or equal to 50% of the Delphi group |

### Standardisation

The selected data elements were mapped to the SNOMED CT terminologies. The June 2022 release of SNOMED CT, which has over 350K active concepts, each with single or multiple synonymous human readable descriptions, was used. These concepts are interrelated in order to provide additional definition or context, with relationships being either hierarchical (eg with “clinical finding” being higher in a hierarchical relationship with “visual acuity”) or non-hierarchical (e.g. ‘‘associated findings”, or with “viral uveitis” having a defining relationship between “is a type” and “infective uveitis”).

In order to further support interoperability, two additional international standards and terminologies were used. Diagnoses were mapped to the International Statistical Classification of Diseases and Related Health Problems, 10th revision (ICD10). Terms were grouped using the Heath Level 7 (HL7) standard "Fast Healthcare Interoperability Resources" (FHIR) generic data structures into Patient, Observation, Condition, Procedure, Medication. (21)

## Results

From the POIG consensus exercise, 160 elements were selected: 89 as high priority items, with 35 as medium and 29 as low priority items. These elements, and response items where appropriate, were grouped into Patient (n= 13 items), Observation (n= 63 items), Condition (n= 20 items), Procedure (n= 44 items), Medication (n= 18 items). SNOMED CT provided IDs which differentiated between concepts such as procedures and findings, and enables anchoring in eye laterality. For example, separate codes were identified for the capture of related but different concepts such as ‘measurement of visual acuity’ (procedure), ‘measured distance visual acuity’ (finding), ‘measured visual acuity in right eye’ (finding), ‘measured visual acuity in left eye’ (finding).

There were four items for which a SNOMED CT ID could not be found: (i) laser photometry or laser flare photometry, a procedure in which light scatter is measured to quantify anterior ocular chamber inflammation; (ii) laser flare photometry results, measured in photos per microseconds; vitreous haze grade, a measure of the degree of inflammatory chance seen in the vitreous cavity of the eye using obscuration of retinal details seen; and (iv) macular thickness as measured using optical coherence tomography. Data elements, response options and associated values can be found via on online repository (DOI: 10.5522/04/24434278).

## Discussion

Through this mapping activity, using international coding and terminologies, we have created a dataset for childhood onset uveitis care and research. This dataset provides a standardised vocabulary for describing clinical concepts, with a semantic interoperability which will support the exchange of data across different systems, organizations, and international or supranational groups.

Clinical research into rare disorders, such as childhood uveitis, faces the common challenge of securing sample sizes sufficiently large as to support robust statistical analysis. This challenge is typically addressed through multi-centre studies.(14) The sharing and harmonisation of data cross different centres is dependent on interoperability of collected datasets. with the aim of facilitating routine and standardized data collection. This is particularly timely for childhood uveitis. Although a rare condition, there is evidence that previous studies have underestimated disease incidence.

Dependent on study setting and year of study, disease incidence estimates now range from 1 to 101 cases per 100,000 children,(6,8) with evidence of rising incidence,(6) and evidence of a higher than expected incidence during and following the COVID-19 pandemic.(22,23) However, the potential benefit of this standardised dataset go beyond this rare disease. The items within the UK’s Royal College of Ophthalmologists recommended dataset for clinical care for adult inflammatory disease(17) are represented within the data items selected for mapping, having been used in the original consensus work undertaken by the Paediatric Ocular Inflammation Group (POIG). This demonstrates the utility of this dataset for adult onset disease. Adult uveitis is one of the commonest reasons for attendance at eye emergency services, and also an important cause of working age blindness.(2) The potential beneficial impact of this dataset on patient care and research is dependent on its use within clinical and research communities. Facilitators for the future implementation of the dataset include the involvement of a national organisation, specifically POIG, in its development, the harmonisation with the RCOphth’s national dataset, the call for such activities from international organisations including the disease specific Multinational Interdisciplinary Working Group for Uveitis in Childhood (MIWGUC),(16) and the broader work underway by the Observational Medical Outcomes Partnership (OMOP).(24–26) Future expansion of the dataset will be needed to ensure coverage of international concepts and care structures.

In summary, this dataset provide a common language for describing clinical information for childhood uveitis. This semantic interoperability will support data re-use, meaningful data storage, and enable accurate and effective communication between healthcare professionals, researchers, and healthcare information technology systems.

## Data Availability

All data produced are available online at DOI: 10.5522/04/24434278

## Acknowledgments

Members of the Paediatric Ocular Inflammation UNICORN Study Group

Professor Jugnoo Rahi, Professor of Ophthalmic Epidemiology, UCL GOS Institute of Child Health, London, UK

Professor Andrew D Dick, Professor of Translational Health Sciences, Bristol Medical School, University of Bristol, Bristol, UK

Professor Alastair Denniston, Professor of Ocular Inflammation, University of Birmingham, UK

Mr Damien C.M.Yeo, Consultant Ophthalmologist, Alder Hey Children’s NHS Foundation Trust, Liverpool

Mr Jose Gonzalez-Martin, Consultant Ophthalmologist, Alder Hey Children’s NHS Foundation Trust, Liverpool

Ms Eibhlin McLoone, Consultant Ophthalmologist, Belfast Royal Victoria Hospital NHS Foundation Trust

Prof Rachel Pilling, Consultant Ophthalmologist, School of Optometry and Vision Science, University of Bradford

Mr John Bradbury, Consultant Ophthalmologist, Bradford Teaching Hospitals NHS Foundation Trust, Bradford Royal Infirmary

Prof Athimalaipet V Ramanan, Consultant Paediatric Rheumatologist, University Hospitals Bristol NHS Foundation Trust

Ms Catherine Guly, Consultant Ophthalmologist, University Hospitals Bristol NHS Foundation Trust

Ms Brinda Muthusamy, Consultant Ophthalmologist, Cambridge University Hospital NHS Foundation Trust

Mr Patrick Watts, Consultant Ophthalmologist, Cardiff and Vale University Health Board Hospitals

Ms Christine Twomey, Clinical Nurse Specialist, Great Ormond Street Hospital NHS Foundation Trust

Ms Reshma Pattani, Specialist Optometrist, Great Ormond Street Hospital NHS Foundation Trust

Mr Clive Edelsten, Consultant Ophthalmologist, Great Ormond Street Hospital NHS Foundation Trust

Mr Daniel Pharoah, Consultant Ophthalmologist, James Paget University Hospitals NHS Foundation Trust

Mr Vernon Long, Leeds Teaching Hospitals NHS Trust, St. James’s University Hospital

Mr Adam Bates, Consultant Ophthalmologist, Maidstone and Tunbridge Wells NHS Trust

Ms Elisabetta Scoppettuolo, Consultant Ophthalmologist, Maidstone and Tunbridge Wells NHS Trust

Prof Jane Ashworth, Consultant Ophthalmologist, Manchester University NHS Foundation Trust

Ms Laura Steeples, Consultant Ophthalmologist, Manchester University NHS Foundation Trust

Mr Harry Petrushkin, Consultant Ophthalmologist, Moorfields Eye Hospital NHS Foundation Trust and Great Ormond Street Hospital NHS Foundation Trust

Ms Dhanes Thomas, Consultant Ophthalmologist, Moorfields Eye Hospital NHS Foundation Trust

Mr Alan John Connor, Consultant Ophthalmologist, Newcastle Upon Tyne Hospitals NHS Foundation Trust

Dr Una O’Colmain, Consultant Ophthalmologist, Ninewells Hospital, Tayside Scotland NHS Board

Mr Anas Injarie, Consultant Ophthalmologist, Norfolk and Norwich University Hospitals NHS Foundation Trust

Mr Narman Puvanachandra, Consultant Ophthalmologist, Norfolk and Norwich University Hospitals NHS Foundation Trust

Ms Archana Pradeep, Consultant Ophthalmologist, Nottingham University Hospitals NHS Trust

Dr Kishore Warrior, Consultant Paediatric Rheumatologist, Nottingham University Hospitals NHS Trust

Ms Srilakshmi Sharma, Consultant Ophthalmologist, Oxford University Hospitals NHS Foundation Trust

Dr Conrad Schmoll, Consultant Ophthalmologist, Princess Alexandra Eye Pavilion / Royal Hospital for Children and Young People, Edinburgh, Lothian NHS Board

Dr Eoghan Millar, Consultant Ophthalmologist, Royal Children’s Hospital, NHS Greater Glasgow and Clyde

Mrs Kate Bush, Consultant Ophthalmologist, Royal Bournemouth Christchurch Hospitals NHS Foundation Trust

Mr M. Ashwin Reddy, Consultant Ophthalmologist, Royal London Hospital, Barts Health NHS Trust

Ms Jessy Choi, Consultant Ophthalmologist, Sheffield Children’s Hospitals NHS Foundation Trust, and Sheffield Teaching Hospitals NHS Foundation Trust

Ms Gisella Cooper, Clinical Nurse Specialist, Sheffield Children’s Hospitals NHS Foundation Trust, and Sheffield Teaching Hospitals NHS Foundation Trust

Ms Kristina May, Consultant Ophthalmologist, University Hospital Southampton NHS Foundation Trust, Southampton General Hospital

Mr Ed Hughes, Consultant Ophthalmologist, Sussex Eye Hospital, University Hospitals Sussex NHS Foundation Trust

Ms Ailsa Ritchie, Consultant Ophthalmologist, St Thomas’ Hospital, Guys and St Thomas’ NHS Trust

The authors would also like to thank the wider Paediatric Ocular Inflammation Group (https://www.ucl.ac.uk/child-health/poig) for their support for the study.

## Author Contributions

All authors contributed to the development of the dataset. ALS and SK mapped the data elements to international terminologies. SK wrote the manuscript. All authors read and approved the final manuscript.

## Availability of data and materials

Data elements, response options and value sets can be accessed via the supplemental documents and institutional repository

## Ethics approval and consent to participate

No permissions were required to access any data used in this study.

